# Identification of two new genetic loci associated with atrial fibrillation in the Taiwanese population-implication of metabolism and fibrosis in atrial fibrillation mechanism

**DOI:** 10.1101/2023.06.15.23291466

**Authors:** Guan-Wei Lee, Jien-Jiun Chen, Sheng-Nan Chang, Fu-Chun Chiu, Pang-Shuo Huang, Eric Y. Chuang, Chia-Ti Tsai

## Abstract

**Background:** Genome-wide association studies (GWASs) have identified common single nucleotide polymorphisms (SNPs) in more than 100 genomic regions associated with atrial fibrillation (AF). Genes for AF identified by GWAS in the Caucasian populations may show ethnic differences in the Asian populations. We sought to identify other novel AF genes in the Taiwanese population by multi-stage GWAS.

**Methods:** In exploratory stage, GWAS with whole genome genotypes (4,512,191 SNPs) were done in 516 young AF Patients (58.1±8.7 years-old, 438 men [84.9%]) from the National Taiwan University AF registry (NTUAFR) and 5160 normal sinus rhythm controls (57.8 ±8.7 years-old, 2460 men [47.7%]) from Taiwan Biobank. Significant loci were replicated in 1002 independent AF patients and 2003 NSR controls, and also in UK biobank (5630 AF cases and 24000 NSR controls). Quantitative trait locus mapping was performed to implicate functional significance.

**Results:** Stage I GWAS revealed 3 loci associated with AF with the genome-wide significance level, which included locus close to previously reported *PITX2* gene (chromosome 4q25, rs2723329, *P*=1.53×10^−10^) and two novel loci close to *RAP1A* and *HNF4G* genes (chromosome 1p13.2, rs7525578, *P*= 1.24×10^−26^; chromosome 8q21.13, rs2980218, *P*=2.19×10^−9^, respectively). They were further validated in a stage II replication population (P=4.60×10^−9^, 4.45×10^−10^ and 6.97×10^−5^ for *RAP1A, PITX2* and *HNF4G*, respectively). These 3 genes were also validated in the UK population. These 3 significant SNPs also show significant association with tissue expressions (*RAP1A* expression in thyroid, *PITX2* in testicular, and *HNF4G* in lymphocyte tissues, respectively).

**Conclusions:** GWAS in Taiwan revealed previously reported *PITX2* and two novel AF genes (*RAP1A* and *HNF4G*) with the most significant locus in *RAP1A*. *RAP1A* and *HNF4G* genes may implicate fibrosis and metabolic pathways, respectively, in the mechanism of AF.

## INTRODUCTION

Atrial fibrillation (AF) is the most common sustained arrhythmia and a major risk factor for stroke, heart failure and cardiovascular death. In the past decade, genome-wide association studies (GWASs) have identified common single nucleotide polymorphisms (SNPs) in more than 100 genomic regions associated with AF, e.g., on chromosomes 4q25 (*PITX2*), 16q22 (*ZFHX3*) and 1q21 (*KCNN3*) ^1–5^. However, these loci do not fully explain the genetic risk for AF, suggesting that additional genetic factors remain to be discovered. Furthermore, several genes for AF identified by GWAS in the Caucasian populations showed ethnic differences in the Asian populations ^6–8^. Our previous GWAS study of copy number variation (CNV) also showed a unique CNV in the *KCNIP1* gene in association with AF in our Taiwanese population^9^. Accordingly, in the present study we sought to identify other novel AF susceptibility SNPs and genes for AF patients in the Taiwanese population.

## METHODS

### Study population

The AF patients were from our cardiovascular clinics in the National Taiwan University AF Registry (NTUAFR). The detailed descriptions for selection of patients have been reported previously ^9–12^. The normal sinus rhythm (NSR) controls were from the general Taiwanese population (Taiwan Biobank [TWB]) ^13, 14^ with whole genome sequencing data for the stage I exploratory population and from the NTUAFR for the stage II validation population. In the TWB, more than 100000 Taiwanese participants had been recruited from the community to facilitate the research of effects of environmental and genetic factors on disease risks and to provide the health information of Taiwanese people.

We used a multi-stage study design with the attempt to minimize false positive findings yet maximize power and efficiency. The study design is shown in **Figure 1**. In the exploratory stage, whole genome GWAS was conducted in 516 young AF patients (mean age 58.1±8.7 years, 271 men [52.5%]) and 5160 NSR controls (57.8±8.7 years, 2460 men [47.7%]). In the replication stage, significant loci in the exploratory stage were validated in 1002 general AF patients and 2003 NSR controls from our cardiovascular clinics (NTUAFR) as in our previous study^9^. All these patients were Taiwanese without population stratification (λ=1.036). The study was approved by the Institutional Review Board (IRB) of the National Taiwan University Hospital (200911002R) and written informed consent was obtained from all the participating individuals.

**Figure 1.**
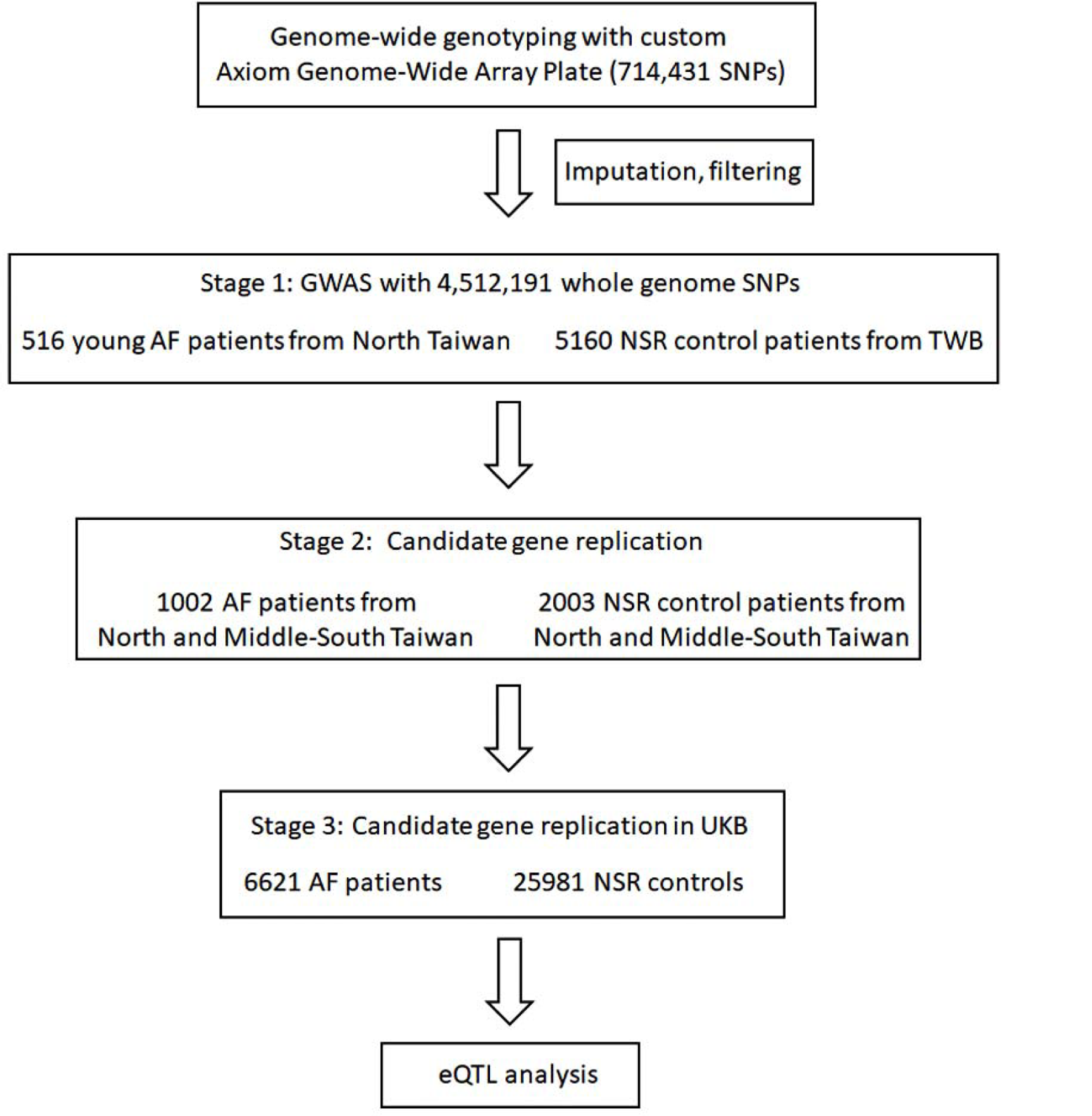
CONSORT flow diagram showing the study design of three-staged genome-wide association study (GWAS). AF, atrial fibrillation; GWAS, genome-wide association study; NSR, normal sinus rhythm; SNP, single nucleotide polymorphism.

### Validation of Taiwan AF genes in the Caucasian population

In the third stage, to more confirm the association of genes identified in the Taiwanese population with AF, we further validated the association in the United Kingdom (UK) population using the UK Biobank genotype data. In brief, UK Biobank is a prospective study of more than 500,000 people living in the UK with long-term follow-up. It was established to allow detailed investigations of the genetic and nongenetic determinants of the diseases of middle and old age ^15^.Participant DNA was genotyped on two arrays, UK BiLEVE and UKB Axiom. Genotypes of ∼800,000 SNPs passing quality controls were imputed to the UK10K reference panel.

Genotypes were called using Affymetrix Power Tools software. Finally, 6621 patients with AF were used as the case population and 25981 patients without AF were used as the control population ^16^.

### Genome-wide genotyping

In the exploratory stage, both the AF patients and NSR controls were genotyped by a custom Axiom Genome-Wide Array Plate that contained 714,431 SNPs, called the TWB chip 2, based on technology developed by Affymetrix. For genotypic quality control, SNPs with a missing call rate > 0.05, Hardy-Weinberg equilibrium (HWE) p-value < 1×10^−6^ in controls, and a minor allele frequency (MAF) < 0.05 were excluded.

### Genotype imputation

Before imputation, we first converted hg38 coordinates to hg19 coordinates (HRC-1000G-check-bim)(https://www.well.ox.ac.uk/∼wrayner/tools/). Then we used Minimac4 for genotype imputation on the 22 chromosomes in the Michigan Imputation Server ^17^, using the 1,000 Genomes Project data of the Han Chinese population (Phase 3 Integrated Release Version 5 Haplotypes) as the reference panel. After imputation, for quality control, we only selected imputed SNPs with imputation quality R^2^ > 0.8, Hardy-Weinberg equilibrium p value > ×10^−6^ and/or MAF ≥ 5%, and complete genotypes in >99% of the control subjects. Finally, 4,512,191 SNPs passed the quality control and were used for downstream GWAS analysis.

### Association analysis

A SNP-based association analysis was performed using PLINK (v1.9) ^18^ based on the binary outcome model of logistic regression. We adjusted the top 10 principal components, age and gender in the association analysis. Manhattan plots and quantile-quantile (Q-Q) plots were generated with the *qqman* package. We used the burden test for rare variant analyses ^19^. Expression quantitative trait loci (eQTL) mapping of novel AF genetic loci was performed by accessing the publicly available database in Genotype-Tissue Expression (GTEx) version 8.

## RESULTS

### Novel AF genetic loci in the Taiwanese population

The baseline characteristics of the study participants are summarized in **Table 1**.

**Table 1.** Patient characteristics of the study population

| Variables | Exploratory population |  | Replication population |  |
| --- | --- | --- | --- | --- |
|  | AF | NSR | AF | NSR |
|  | (n=516) | (n=5160) | (n=1002) | (n=2003) |
| Age, mean±SD | 58.1±8.7 | 57.8±8.7 | 68.7±12.1 * | 60.7±14.2 |
| BMI, mean±SD | 25.3±3.8 * | 24.4±3.6 | 24.4±4.6 | 24.8±4.0 |
| Female, n (%) | 245 (47.5) | 2695 (52.3) | 409(40.8) | 916 (45.7) |
| Male, n (%) | 271 (52.5) | 2460 (47.7) | 593 (59.2) | 1087 (54.3) |
| Diabetes, n (%) | 18 (12.0) * | 458 (8.9) | 238 (23.8) * | 327 (16.3) |
| Hyperlipidemia, n (%) | 25 (16.7) | 668 (13.0) | 134 (13.4) | 278 (13.9) |
| Hypertension, n (%) | 50 (33.3) * | 1104 (21.4) | 576 (57.5) * | 819 (40.9) |
| CAD, n (%) | 8 (5.3) * | 150 (2.9) | 246 (24.6) | 402 (20.0) |

AF, atrial fibrillation; BMI, body mass index; CAD, coronary artery disease; NSR, normal sinus rhythm.
\* P <0.05 compared to NSR.

As expected, the mean age and the percentages of hypertension, coronary artery disease, diabetes or stroke in the AF patients were higher than those in the NSR controls both in the exploratory and validation populations.

GWAS was performed in the stage I subjects, using the Axiom Genome-Wide Array Chip (714,431 SNPs)**(Figure 2)**. After quality control filtering and imputation, there was a total of 4,512,191 SNPs. The Manhattan plot of GWAS is shown in **Figure 2A** and QQ plot in **Figure 2A**. Three loci were associated with AF with *P* < 5 × 10^−8^ in the exploratory stage **(Figure 2A)**. While the *PITX2* locus was previously identified as the most significant AF locus ^1–5^, two novel genes (*RAP1A*, and *HNF4G*) were significantly associated with AF in the Taiwanese population. The most significant association was shown in 1p13.2/ *RAP1A* rs7525578 (odds ratio [OR] 2.74, 95% confidence interval [CI] 2.28–3.30; *P =*1.24×10^−26^), followed by 4q25/*PITX2* rs2723329 (OR 1.54, 95% CI 1.35-1.76, *P*=2.19×10^−9^) and then 8q21.13/*HNF4G* rs2980218 (OR 1.49, 95% CI 1.31-1.70, *P*=2.19×10^−9^). **Figure 3** shows the regional plots for these three significant AF loci.

**Figure 2.**
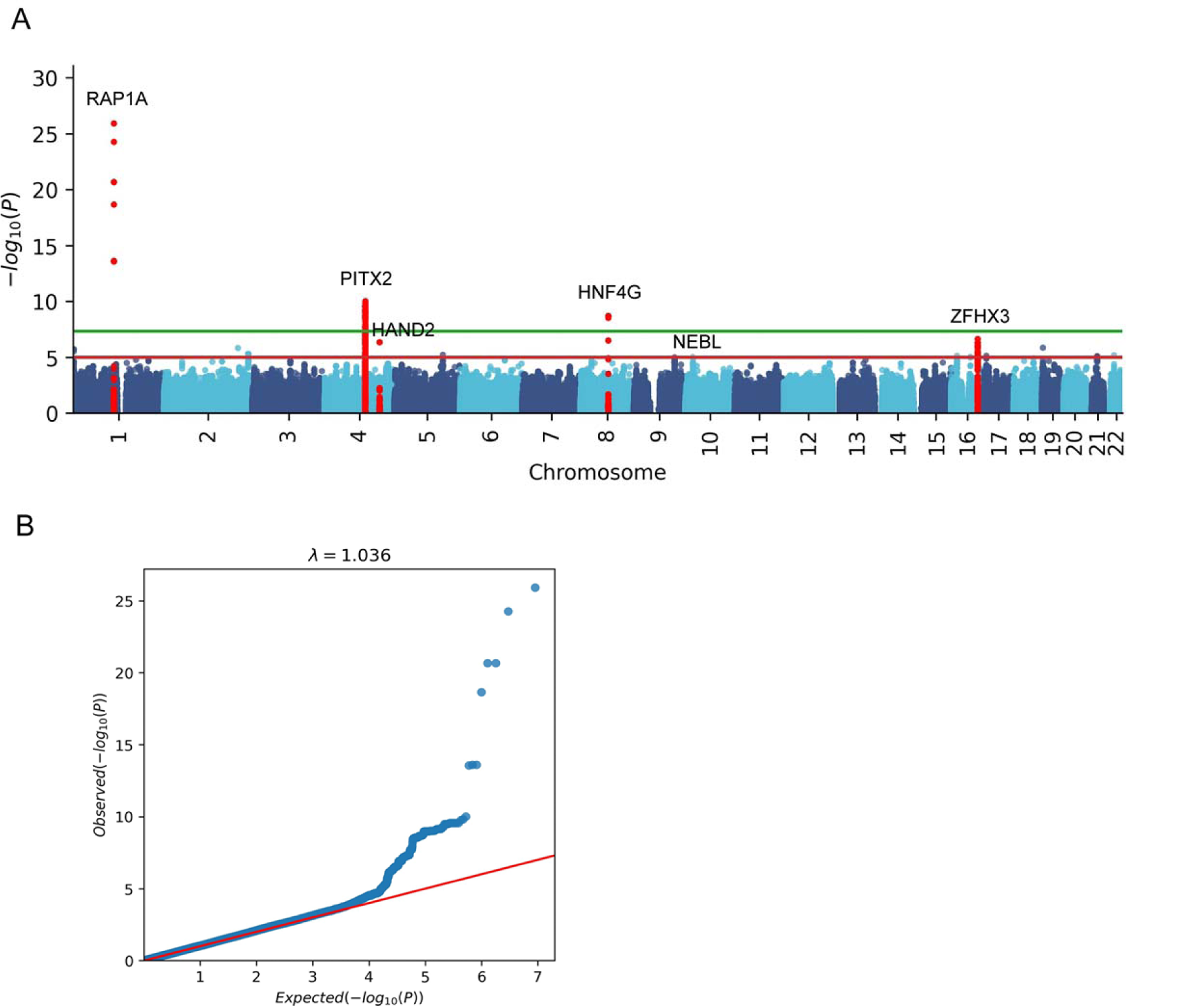
Manhattan and QQ plots of genome-wide association study (GWAS) results. A, Three loci/genes (*RAP1A, PITX2* and *HNF4G*) show associations with AF with *P* < 5 × 10^−8^ (above the green line) and two loci/genes (*HAND2* and *ZFHX3*) show associations with AF with *P* < 10^−5^ (above the red line) in the exploratory stage. B, QQ-plot of the GWAS scan. Most *P*-values were similar to the expected diagonal in the QQ-plot, which indicates the appropriateness of the GWAS model (lambda = 1.036).

**Figure 3.**
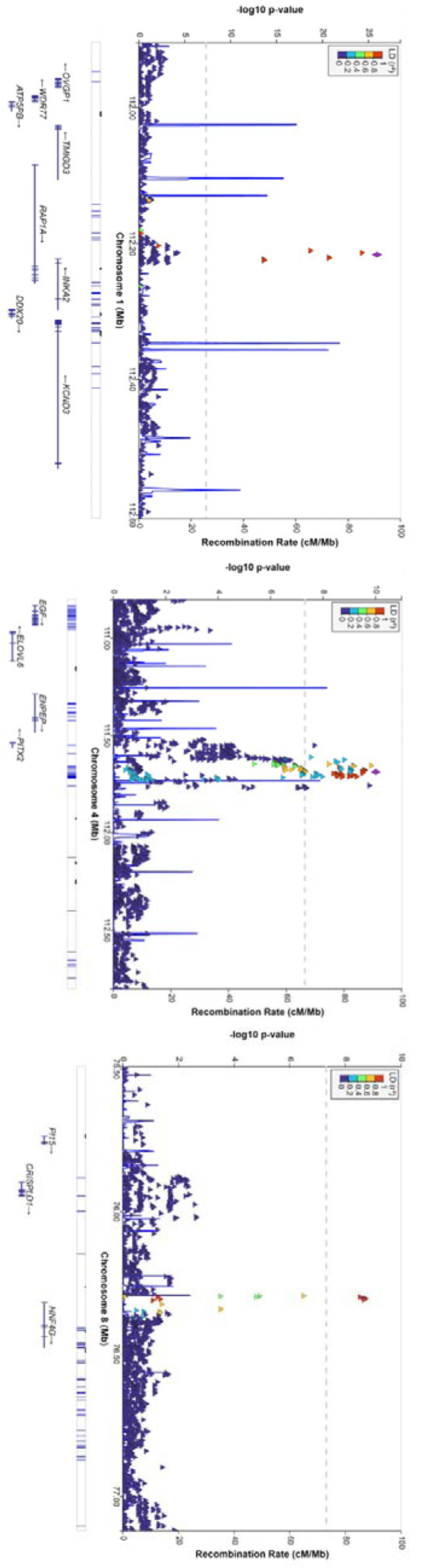
Regional plots of variants that how associations with AF with *P* < 5 × 10^−8^. Regional plots of SNPs centered on the lead SNP around the *RAP1A* gene (upper panel, *PITX2* gene (middle panel) and *HNF4G* gene (lower panel) are shown. For each SNP, the chromosomal location is shown on the x-axis and the significance level for association with AF is indicated by a –log10P value on the y-axis. P-values are expressed as –log10(P)(y-axis) for every tested SNP ordered by chromosomal location (x-axis).

### Validation of GWAS AF genes in a replication population

The significant loci identified in the exploratory GWAS were further validated in a replication population with 1002 AF patients and 2003 NSR controls. The association with AF was replicated in all these 3 genes. The ORs and *P* values were 1.72 (95% CI 1.42-2.07), 1.41 (95% CI 1.27-1.58) and 1.25 (95% CI 1.12-1.40), and 4.60×10^−9^, 4.45×10^−10^ and 6.97×10^−5^ for *RAP1A*, *PITX2* and *HNF4G*, respectively.

### Validation of Taiwan AF genes in the UK population

We further validated the association of *PITX2, RAP1A* and *HNF4G* with AF in the UK Biobank. These 3 genes were also significantly associated with AF in the UK population. The *P* value for *PITX2* gene (rs200657990), *RAP1A* gene (rs141883790) and *HNF4G* gene (rs114415293) were 2.93 ×10^−43^, 0.0015 and 0.0022, respectively.

### Validation of previous AF genes in our Taiwanese population

A total of more than 100 genetic loci have been shown to be associated with AF in the AF multi-ethnic meta-analyses ^5^. We also validated all these loci in our Taiwanese population and the results are shown in **Table 2**. All the genes with *P* values less than 0.1 were listed. The *P* value for significant association was corrected by the Bonferroni method, and a P less than 0.05/100 were considered significant.

**Table 2.** Replication of known AF genes in the Taiwanese population

| #Chr | Position | SNP | Gene | Allele | OR | 95% CI | P |
| --- | --- | --- | --- | --- | --- | --- | --- |
| 1 | 22282619 | rs7529220 | HSPG2 | C/T | 0.772 | 0.655-0.910 | 0.0020505 |
| 1 | 112392360 | rs12044963 | KCND3 | G/T | 1.164 | 1.020-1.328 | 0.0242902 |
| 1 | 116297758 | rs4073778 | CASQ2 | C/A | 1.203 | 1.051-1.377 | 0.0072332 |
| 1 | 170591310 | rs651386 | PRRX1 | A/T | 0.860 | 0.751-0.984 | 0.0282370 |
| 1 | 203026591 | rs17461925 | PPFIA4 | A/G | 0.803 | 0.680-0.949 | 0.0100012 |
| 1 | 205691316 | rs4951258 | NUCKS1 | G/A | 1.149 | 0.999-1.321 | 0.0517686 |
| 3 | 38767315 | rs6801957 | SCN10A | C/T | 0.866 | 0.735-1.02 | 0.0856507 |
| 3 | 179170494 | rs4855075 | GNB4 | C/T | 1.220 | 1.023-1.454 | 0.0266717 |
| 4 | 111658394 | rs75021220 | PITX2 | C/T | 1.519 | 1.330-1.736 | 7.22E-10 |
| 4 | 174448143 | rs78164752 | HAND2 | T/G | 1.434 | 1.248-1.702 | 4.92E-06 |
| 4 | 174642789 | rs74500426 | HAND2-AS1 | G/T | 0.809 | 0.649-1.009 | 0.0598078 |
| 5 | 113736416 | rs716845 | KCNN2 | G/A | 1.223 | 1.009-1.482 | 0.0401149 |
| 7 | 150661409 | rs7789146 | KCNH2 | G/A | 1.119 | 0.980-1.278 | 0.0965402 |
| 8 | 141746324 | rs4355822 | PTK2 | A/G | 0.835 | 0.727-0.960 | 0.0110472 |
| 10 | 21157621 | rs2296610 | NEBL | G/T | 1.349 | 1.144-1.591 | 0.0003795 |
| 10 | 75414344 | rs60212594 | SYNPO2L | G/C | 0.860 | 0.727-1.016 | 0.0754544 |
| 10 | 105342672 | rs11598047 | NEURL1 | A/G | 1.178 | 1.00-1.387 | 0.0500383 |
| 14 | 32981484 | rs2145587 | AKAP6 | G/A | 1.277 | 1.121-1.455 | 0.0002357 |
| 14 | 73361270 | rs3814864 | DPF3 | A/G | 1.122 | 0.986-1.276 | 0.0813060 |
| 16 | 73051620 | rs2106261 | ZFHX3 | C/T | 1.421 | 1.244-1.624 | 2.42E-07 |
| 17 | 44019712 | rs242557 | MAPT | A/G | 1.141 | 1.001-1.299 | 0.0478401 |

AF, atrial fibrillation; #Chr, chromosome number; CI, confidence interval; OR, odds ratio; SNP, single nucleotide polymorphism.

Among these 100 previously defined AF genes, five genetic loci were reproducibly associated with AF with statistical significance in our Taiwanese population: *PITX2* (rs75021220), *ZFHX3* (rs2106261), *HAND2* (rs78164752), *AKAP6* (rs2145587), and *NEBL* (rs2296610). The most significant association was in the *PITX2 gene* (rs75021220)(odds ratio [OR] 1.52; 95% confidence interval (CI) 1.33–1.73, *P =* 7.22 × 10^−10^], followed by *ZHFX* (rs2106261)(OR 1.42; 95% CI 1.24–1.62, *P =* 2.42 × 10^−7^). Interestingly, the association of *HAND2* is similar to the previous finding in a Korean population in the effect size (OR 1.43; 95% CI 1.20–1.70, *P =* 4.92 × 10^−6^) ^8^.

### Expression quantitative trait loci (eQTL) mapping of novel AF genetic loci

The effect of novel susceptibility SNPs on expression of genes in various human tissues was evaluated by investigating eQTLs. Several significant associations between SNPs in novel susceptibility genes and gene expression were found. The top AF SNP in the *RAP1A* gene (rs7525578) was significantly associated with *RAP1A* expression in the thyroid tissue (*P*= 8.02 × 10^−8^). Two other AF risk SNPs at the same locus also had a significant eQTL association with *RAP1A* expression in the thyroid tissue (rs12141323, *P*=6.12 × 10^−8^; rs6704009, *P*=8.34 × 10^−8^). Among SNPs associated with AF in the *PITX2* gene, rs2723329 had a significant eQTL association with *PITX2* expression in the testis (*P* = 1.07 × 10^−4^). The most significant SNP in the *HNF4G* gene (rs2926606) was significantly associated with *HNF4G* expression in lymphocytes (*P* =1.24 × 10^−6^). The other SNP at the same locus also had a significant eQTL association with *HNF4G* expression in lymphocytes (rs2980231, *P*=2.50× 10^−5^).

### Rare variant analysis

We used burden test to search possible rare variants among the 23000 human genes that might be associated with AF in our Taiwanese population. We found only one gene (*UGT2B17*) reaching the genome-wide significant level (0.05/23000)(**Figure 4**). However, the association of rare variants in *UGT2B17* was not replicated in the replication population. This result is similar to a recent rare variant analysis of AF in the Caucasian population, also showing no rare variant significantly associated with AF^20^.

**Figure 4.**
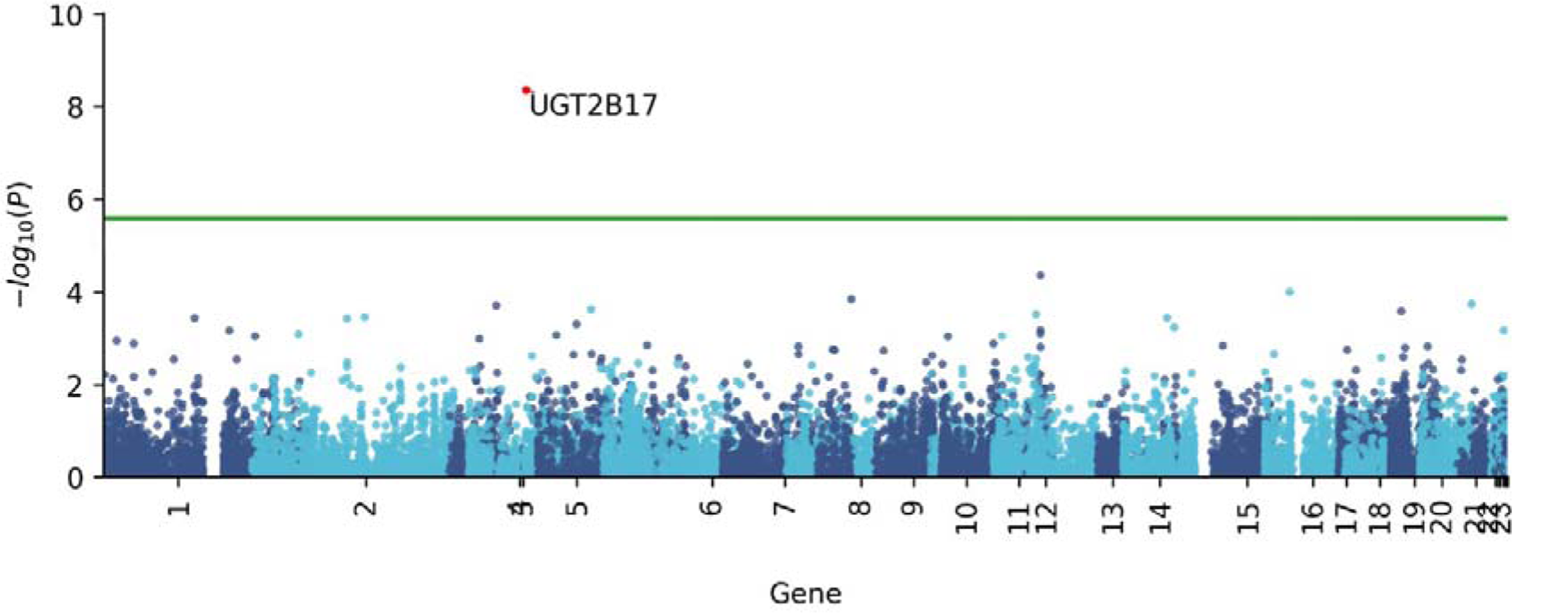
Manhattan plot for each gene obtained from the gene-based burden test. Green line demarcates the Bonferroni significance threshold (p < 0.5/20,000).

## DISCUSSION

In the present study, we found two novel genes associated with AF specifically in the Taiwanese population, which had never been reported before our study.

In addition to Japanese and Korean AF GWASs, our study is the third AF GWAS in the Asia-Pacific region. Our case number was close to the Korean AF GWAS population. In all these 3 AF GWAS studies in the Asia-Pacific region, we all showed the *PITX2* gene highly significantly associated with AF. In the present Taiwan AF GWAS, we identified two novel genes at the genome-wide significant level (5×10^−8^), among them *RAP1A* was even more significant than the *PITX2* gene. The association of these two novel genes with AF were further validated in the UK population.

Therefore, we continued to add new AF genes to the AF GWAS catalog (*HNF4G* and *RAP1A*). In the Japanese AF GWAS studies, 6 new genes (*KCND3, PPFIA4, SLC1A4-CEP68, HAND2, NEBL* and *SH3PXD2A* genes) were added into the AF gene group ^7^. Among these 6 novel AF genes in the Japanese population, *PPFIA4* and *HAND2* were also validated in the Korean AF GWAS ^8^. In our Taiwanese GWAS, the *HAND2* gene was also significantly associated with AF with a more conservative *P* value threshold (*P* <10^−5^). Our case number was close to the Korean AF GWAS but revealed not as many previous AF loci as the Korean study. This may be due to chance or different genetic architecture of AF in different populations.

What is the plausible explanation for the association of the *HNF4G* gene with AF? *HNF4G* also showed significant QTL effect in the present study. Recently, *HNF4G* has been reported as a risk gene for obesity ^21^. Obesity has been reported to be an important risk factor of AF^22, 23^. Importantly, single weight reduction could decrease the burden of AF^24^. In our AF GWAS population, AF patients had a significantly higher mean BMI compared with that of the NSR controls. It is possible that the association of the *HNF4G* gene with AF is body-weight dependent.

However, in our replication population, AF patients did not have a significantly higher mean BMI compared with that of the NSR controls and we did find a very significant association of *HNF4G* gene with AF. It has also been demonstrated that an obesity gene could be a disease-causing gene for premature coronary artery disease independent of BMI ^25^. Therefore, it is also possible that *HNF4G* is related to AF mechanism through metabolic pathways independent of BMI or obesity^26, 27^.

*RAP1A* gene encodes for Rap1 GTPase. Data have pointed to a significant role of Rap1 GTPase in reducing fibrotic gene expression and myofibroblast activation suggesting possible new therapeutic strategies to combat the damaging effects of cardiac fibrosis ^28^. It had also been shown that Rap1 GTPase controlled expression of fibrosis and stress related genes in cardiac fibroblasts isolated from type 2 diabetic mice.

It is possible that the role of *RAP1A* in AF mechanism may be amplified in the scenario of diabetes or metabolic disorder, and *RAP1A* and *HNF4G* may interaction with each other to cause AF. This might explain why both *RAP1A* and *HNF4G* were identified as AF risk genes in our study, linking metabolism and fibrosis to the mechanism of AF^29, 30^. It has been well established that atrial fibrosis plays a pivotal role in the AF mechanism ^31–33^. Therefore, *RAP1A* and *HNF4G* may present a risk gene set of cardiac fibrosis in metabolic disorders or obesity.

eQTL analysis has been established to provide information regarding whether a specific SNP is associated with gene expression ^34^. Integration of GWAS and eQTL can help us to know the possible mechanism why a genetic association causes the disease ^35, 36^. The GTEx project provides a comprehensive publicly accessible database linking the whole-genome and transcriptome sequencing data and whole genome genotype data using 53 normal human tissues from nearly 1,000 individuals to address the effects of functional variations.

In our eQTL analyses, we found SNPs in all the 3 significant genes were associated with gene expression in various tissues. Of note, the significant SNP in the *HNF4G* gene was highly associated with *HNF4G* expression in lymphocytes. This may indicate a pivotal role of inflammation in metabolism and AF. There has been much evidence showing that obesity or metabolic dysregulation predisposes to cardiovascular diseases through inflammation^37, 38^.

There are limitations in the present study. First, we did not directly prove that *RAP1A* and *HNF4G* are involved in the mechanism of AF. Further studies using knockout mice may be performed to prove the link of *RAP1A* gene to atrial fibrosis and *HNF4G* to metabolism or obesity related inflammatory pathways in AF mechanism. Second, although the association of the novel *RAP1A* and *HNF4G* genes with AF was also validated in the UK population, the strength of association was not as large as in our Taiwanese population. There still exists racial difference in the genetic architecture of AF. Third, the case number might be too low for the rare variant analyses. Finally, in the eQTL analyses, although it has been well known that *PITX2* and *RAP1A* are expressed in the left atrium, the significant SNPs of these two genes are associated with tissue expression in testis and thyroid, respectively. These are only database derived analyses and real analyses in the human left atrium are warranted.

## Data availability Statement

The raw data supporting the conclusions of this article will be made available by the authors, without undue reservation.

